# Robust SARS-CoV-2 antibody and T cell immunity following three COVID-19 vaccine doses in inflammatory bowel disease patients receiving anti-TNF or alternative treatments

**DOI:** 10.1101/2022.12.13.22283434

**Authors:** Eva Zhang, Thi H O Nguyen, Lilith F Allen, Lukasz Kedzierski, Louise C Rowntree, So Young Chang, Wuji Zhang, Jennifer R Habel, Isabelle J Foo, Tejas Menon, Jeni Mitchell, Rupert W Leong, Katherine A Bond, Deborah A Williamson, Katherine Kedzierska, Britt Christensen

**Author notes:** Email correspondence to Associate Professor Britt Christensen and Professor Katherine Kedzierska. Authors contributed equally to this study (Eva Zhang, Thi H O Nguyen). Equal senior authors (Katherine Kedzierska, Britt Christensen). **Authorship Contributions** Eva Zhang, BMed MD (Conceptualisation: supporting; Data curation: equal; Formal analysis: equal; Investigation: Equal; Methodology: Supporting; Project administration: Lead; Writing – original draft: Equal; Writing – review & editing: Supporting) Thi O N Nguyen, PhD (Conceptualisation: supporting; Data curation: supporting; Formal analysis: equal; Investigation: Equal; Methodology: Supporting; Resources: Supporting; Writing – original draft: Equal; Writing – review & editing: Supporting) Lilith Allen, MS (Data curation: supporting; Formal analysis: supporting; Investigation: supporting; Methodology: Supporting; Writing – review & editing: Supporting) Lukasz Kedzierski, PhD (Formal analysis: supporting; Investigation: supporting; Methodology: Supporting; Writing – review & editing: Supporting) Louise Rowntree, PhD (Formal analysis: supporting; Investigation: supporting; Writing – review & editing: Supporting) So Young Chang, MS (Formal analysis: supporting; Investigation: supporting) Wuji Zhang, MS (Formal analysis: supporting; Investigation: supporting) Jennifer R Habel, MS (Formal analysis: supporting; Investigation: supporting) Isabelle J Foo, MS MS (Formal analysis: supporting; Investigation: supporting) Tejas Menon, MS MS (Formal analysis: supporting; Investigation: supporting) Jeni Mitchell (Project administration – supporting) Rupert W Leong (Supervision – supporting, Writing – review & editing: Supporting) Katherine A Bond (Conceptualisation: supporting; Data curation: supporting; Investigation: supporting; Resources: Supporting; Writing – review & editing: Supporting) Deborah A Williamson (Formal analysis: supporting; Investigation: supporting; Methodology: Supporting; Writing – review & editing: Supporting) Katherine Kedzierska (Conceptualization: Equal; Funding acquisition: Lead; Investigation: Supporting; Methodology: Supporting; Project administration: Supporting; Supervision: Equal; Visualization: Supporting; Writing – review & editing: Equal). Britt Christensen, MBBS PhD (Conceptualization: Equal; Funding acquisition: Supporting; Investigation: Supporting; Methodology: Supporting; Project administration: Supporting; Supervision: Equal; Visualization: Supporting; Writing – review & editing: Equal). **Contributions** BC, KK and RWL supervised the study. EZ, THON, LFA, LK, KAB, DAW, BC and KK designed the experiments. THON, LFA, LK and LCR performed humoral and T cell experiments. THON, LFA, LCR, SYC, WZ, JRH and IJF processed bloods and performed whole blood staining. EZ, THON, LFA, LK, TM and KAB analysed data. EZ, JM and BC recruited the IBD vaccinated patients and collated patients’ clinical data. LCR, JM, KAB and DAW recruited the healthy vaccinated cohorts. EZ, THON, LFA, BC and KK wrote the manuscript. All authors reviewed and approved the manuscript. **Competing interests** None declared.

## Abstract

**BACKGROUND AND AIMS:** Vaccine-mediated immune responses in patients with inflammatory bowel disease (IBD) may be influenced by IBD therapies. We investigated in-depth humoral and T-cell responses to SARS-CoV-2 vaccination in IBD patients following three COVID-19 vaccine doses.

**METHODS:** Immune responses of 100 SARS-CoV-2-uninfected IBD patients on varying treatments were compared to healthy controls (n=35). Anti-S1/2 and anti-RBD SARS-CoV-2-specific antibodies, CD4^+^ and CD8^+^ T-cell responses were measured at baseline and at five time-points after COVID-19 vaccination.

**RESULTS:** Anti-S1/2 and anti-RBD antibody concentrations at ∼1 month after second dose vaccination were significantly lower in anti-TNF-treated patients compared to non-TNF IBD patients and healthy controls (126.4 vs 262.1 and 295.5, p<0.0001). Anti-S1/2 antibodies remained reduced in anti-TNF treated patients before and after the third dose (285.7 vs 365.3, *p*=0.03), although anti-RBD antibodies reached comparable titres to non-TNF patients. Anti-RBD antibodies were higher in the vedolizumab group than controls after second dose (4.2 vs 3.6, p=0.003). Anti-TNF monotherapy was associated with increased CD4^+^ and CD8^+^ T-cell activation compared to combination anti-TNF patients after second dose, but comparable after third dose. Overall, IBD patients demonstrated similar CD4^+^/CD8^+^ T-cell responses compared to healthy controls regardless of treatment regimen.

**CONCLUSIONS:** Anti-TNFs impaired antibody concentrations when compared to non-TNF patients and controls after two vaccine doses. These differences were not observed after the third vaccine dose. However, vaccine induced SARS-CoV-2-specific T cell responses are robust in anti-TNF-treated patients. Our study supports the need for timely booster vaccination particularly in anti-TNF treated patients to minimise the risk of severe SARS-CoV-2 infection.

## INTRODUCTION

The SARS-CoV-2 pandemic has had devastating and far-reaching impacts worldwide, prompting rapid research and development into vaccines in the hope of mitigating its impact. The rapidity of vaccine development and relative paucity of experience has created a significant challenge. This is particularly the case for groups excluded from vaccine trial populations, such as immunosuppressed individuals including those with inflammatory bowel disease (IBD) ^1,2^.

Most published data in IBD patients pertain to SARS-CoV-2 antibody responses, which demonstrate that antibody concentrations can be impaired by immune modifying medications. Anti-TNF therapy impairs peak antibody concentrations achieved after 2 doses of either mRNA or viral vector vaccines ^3–6^ compared to vedolizumab, ustekinumab and thiopurine monotherapy. Antibody concentrations also decline more quickly in infliximab-treated patients, although reassuringly, most IBD patients seroconverted after two doses regardless of immunosuppressive regime ^5^. There was no association between trough anti-TNF level and antibody titres ^7^. Tofacitinib has also been associated with a lower antibody response in the setting of mRNA and viral vector vaccines ^8^. Older age and thiopurine use have also been found to impair antibody responses.

Antibody concentrations wane with time after two-dose vaccination which exposes individuals to breakthrough infection ^9^. A third mRNA vaccine dose leads to a rise in antibody levels ^10^ across all medication regimes in those with IBD. A third vaccine dose also leads to seroconversion in those who fail to seroconvert after two doses ^10,11^. However, antibody concentrations remain attenuated with anti-TNF therapy after third dose vaccination compared to non-TNF treated patients ^8,11,12^. Furthermore, the risk of breakthrough COVID-19 infection is increased in anti-TNF-treated individuals compared to those receiving vedolizumab. This risk bears no correlation to anti-spike antibody concentrations, which is a reflection of viral escape in the setting of mutating viral variants ^12^.

Although antibodies have been the primary measure of vaccine efficacy thus far, T cell responses are another key immunological parameter. The few studies examining this in IBD have shown that the T cell response is decoupled from the antibody response ^3,13^. T cell responses appear to be robust amongst IBD patients and comparable to controls ^14,15 16^, with the exception of tofacitinib ^8^. Interestingly, spike-stimulated PBMCs from anti-TNF-treated patients demonstrated higher levels of IFN-γ production than controls both at 3 and 6 months after two-dose vaccination, while spike-stimulated whole-blood supernatant produced a significantly higher IL-10 response in anti-TNF^+^ patients ^16^. An analysis of 303 IBD patients further demonstrated a positive association with anti-TNF and augmented T Cell Receptor (TCR) clonal depth ^13^.

Vaccine-induced immunity is complex, particularly in the setting of immunosuppression and emerging virus variants. In our study, we sought to clarify the utility of third dose vaccination and the T cell response in IBD patients, which is poorly defined thus far.

## METHODS

### Subject recruitment

Participants with IBD were recruited prospectively at the Royal Melbourne Hospital (Melbourne, Australia) with approval from Melbourne Health (HREC/74403/MH-2021). Patients were aged 16 years and over with a diagnosis of IBD and on a stable maintenance medication regimen (5ASA, anti-TNF, anti-metabolite, IL12/13 antagonists, vedolizumab, or tofacitinib) for over 2 months. Healthcare workers who did not have IBD and were not on immunosuppressive medication were enrolled as controls with approvals from Melbourne Health (HREC/68355/MH-2020) and University of Melbourne (HREC 22268, 21626). All participants provided written informed consent. No participants had a clinical history of SARS-CoV-2 infection at enrolment. Variables captured included age, sex, comorbidities, height and weight, smoking status and type and date of vaccination.

#### Patient involvement

Participants were not involved in the study design, nor outcome measures. Upon publication findings will be disseminated to participants via a newsletter.

### Sample collection

Blood collection was undertaken at 4 time points: at baseline (V0), after dose 1 (V1); after dose 2 (V2); and ∼1 month post dose 2 (V3). Blood collection was also undertaken at 2 further time points: from 30 days before dose 3 (V5) and from ∼1-3 months after dose 3 (V6). Out of 100 total patients, 88 were collected from V0-V3+/- V5-V6, and 12 joined the study prior to the third dose where samples were only collected for V5-V6. Whole blood was collected in sodium heparin tubes and peripheral blood mononuclear cells (PBMCs) were isolated via Ficoll–Paque separation, cryopreserved in 10% DMSO/FCS and stored in liquid nitrogen. PMBC isolation was performed within 24 hours of venepuncture.

### SARS-CoV-2-specific antibodies

Spike protein S1/2 receptor binding domain specific IgG antibodies were measured using the LIAISON DiaSorin electrochemiluminenscence immunoassay. At study entry and all subsequent time points, all participants were tested for possible recent SARS-CoV-2 infection using the Abbott anti-nucleocapsid protein IgG immunoassay. Index values below 1.4 were considered negative and at which participants were deemed to have no evidence of prior infection. IgG ELISA was used to assess the presence of antibodies against ancestral SARS-CoV-2 RBD in plasma, as previously described ^17^. RBD seropositivity was defined as 2 standard deviations above the mean of the cohort at V0.

### Immune cell activation in whole blood

Whole blood staining of V0-V3 timepoints was performed essentially as previously described^18^.

### SARS-CoV-2-specific T cells

Activation induced maker (AIM) Assay was performed on PBMCs at V0, V3 and V6 timepoints according to Grifoni *et al*. with minor modifications ^19^. PBMCs were plated at 1×10^6^ per well and stimulated with 10μg/ml SARS-CoV-2 Spike peptide pool (181 peptides, 0.06μg/ml per peptide, BEI Resources, NIAID, NIH) or DMSO and cultured at 37 °C/5% CO_2_ for 24 hours before being stained with a panel of cellular surface markers [CXCR5-BV421 (562747; BD Biosciences), CD3-BV510 (317332; BioLegend), CD8-BV605 (564116; BD Biosciences), CD4-BV650 (563875; BD Biosciences), CD25-BV711 (563159; BD Biosciences), CXCR3-BV786 (353738; BD Biosciences), CD137-APC (309810; BioLegend), CD27-AF700 (560611; BD Biosciences), CD14/CD19-APC-H7 (560180/560252; BD Biosciences), Live/Dead NIR (L34976; Invitrogen), CD69-PerCPCy5.5 (310925; BioLegend), CD134-PE (340420; BD Biosciences), CD95-PE-CF594 (562395; BD Biosciences), CD45RA-PeCy7 (337167; BD Biosciences)]. Cells were then fixed with 1% PFA before acquisition on a LSRII Fortessa (BD). Activated CD4^+^ T cells were defined as CD137^+^CD134^+^ whilst activated CD8^+^ T cells were defined as CD137^+^CD69^+^.

### Statistical analyses

Anti-SARS-CoV-2 spike protein and receptor binding domain antibody concentrations are reported as geometric means and SD. Other continuous data are reported as median ± interquartile range, and discrete data as numbers and percentages. Log transformed concentrations of anti-SARS-CoV-2 spike protein antibodies were compared between different groups by independent t-tests and ANOVA. Mann-Whitney U-test (unpaired) and Wilcoxin signed-rank test (paired) were used for comparisons between two groups. Kruskal-Wallis test with Dunn’s multiple comparisons was used to compare more than two groups. Spearman’s correlation (r_s_) was calculated to determine the correlation between antibody and T cell responses. Statistical analyses were performed with GraphPad Prism v9 and IBM SPSS Statistics v26. All tests were two-tailed and p values of less than 0.05 were considered significant.

## RESULTS

### IBD patients on varying treatment regimens generate robust antibody responses following SARS-CoV-2 vaccination

The study included 100 IBD participants recruited between July to December 2021, compared to 35 healthy controls. Participant characteristics are outlined in table 1. 30 participants with ulcerative colitis (UC) or indeterminate colitis, and 70 with Crohns disease (CD) were taking various medications: 9 5ASA, 3 thiopurine/methotrexate monotherapy, 15 anti-TNF monotherapy, 40 anti-TNF combination therapy with immunomodulator, 18 IL-12/IL-23 (ustekinumab), 14 vedolizumab, 1 tofacitinib. 89 patients received BNT162b2 (Pfizer–BioNTech), and 11 received ChAdOx1 nCoV-19 (Oxford–AstraZeneca) (figure 1A). All participants had no evidence of prior SARS-CoV-2 infection at enrolment. During the study, 4 IBD patients were infected with SARS-CoV-2 after the first (n=1) and third dose (n=3), based on presence of anti-nucleocapsid antibodies, of which their datapoints after infection were removed from analyses.

**Table 1:**
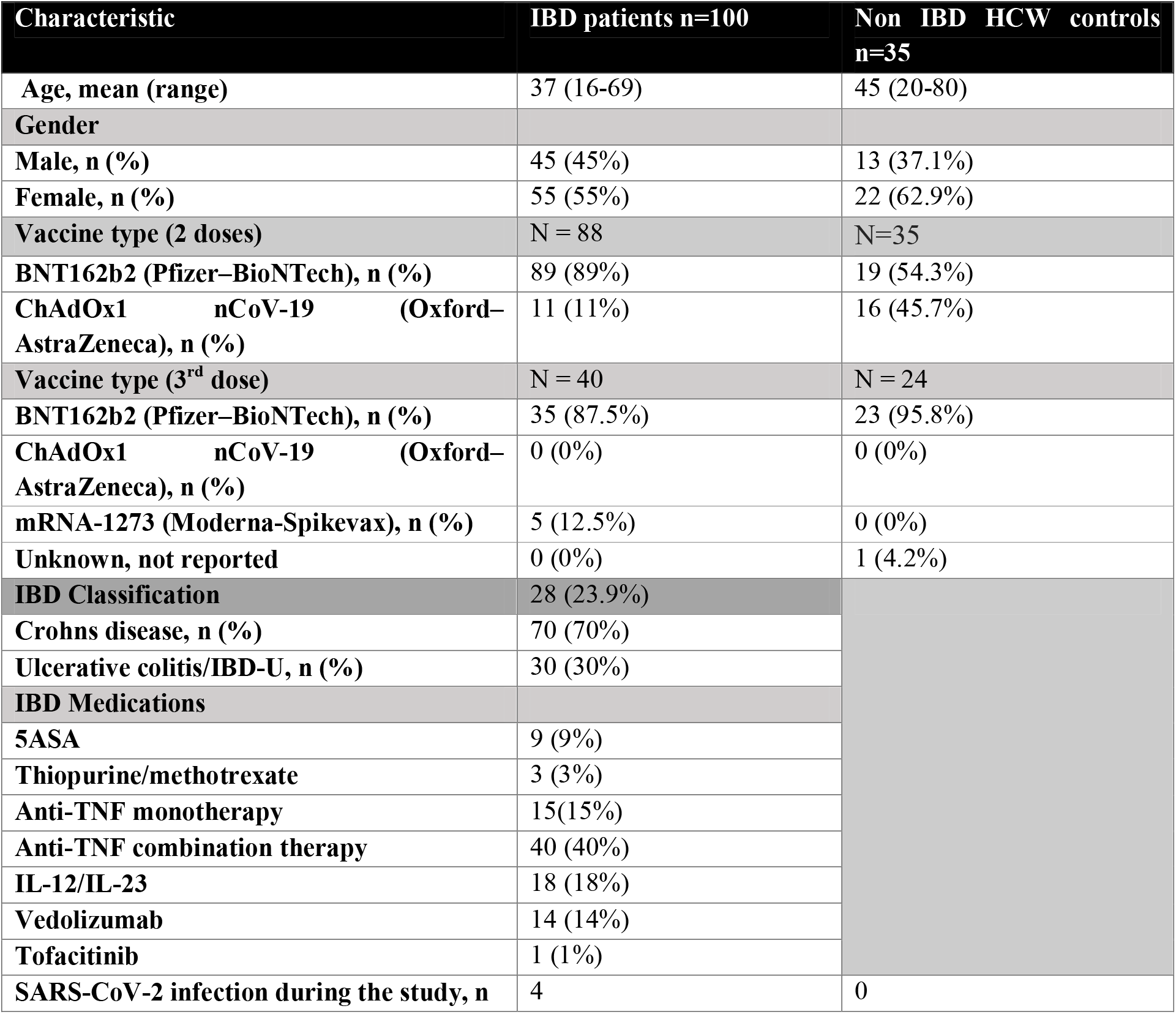
Characteristics of IBD patients and healthy controls

**Figure 1.**
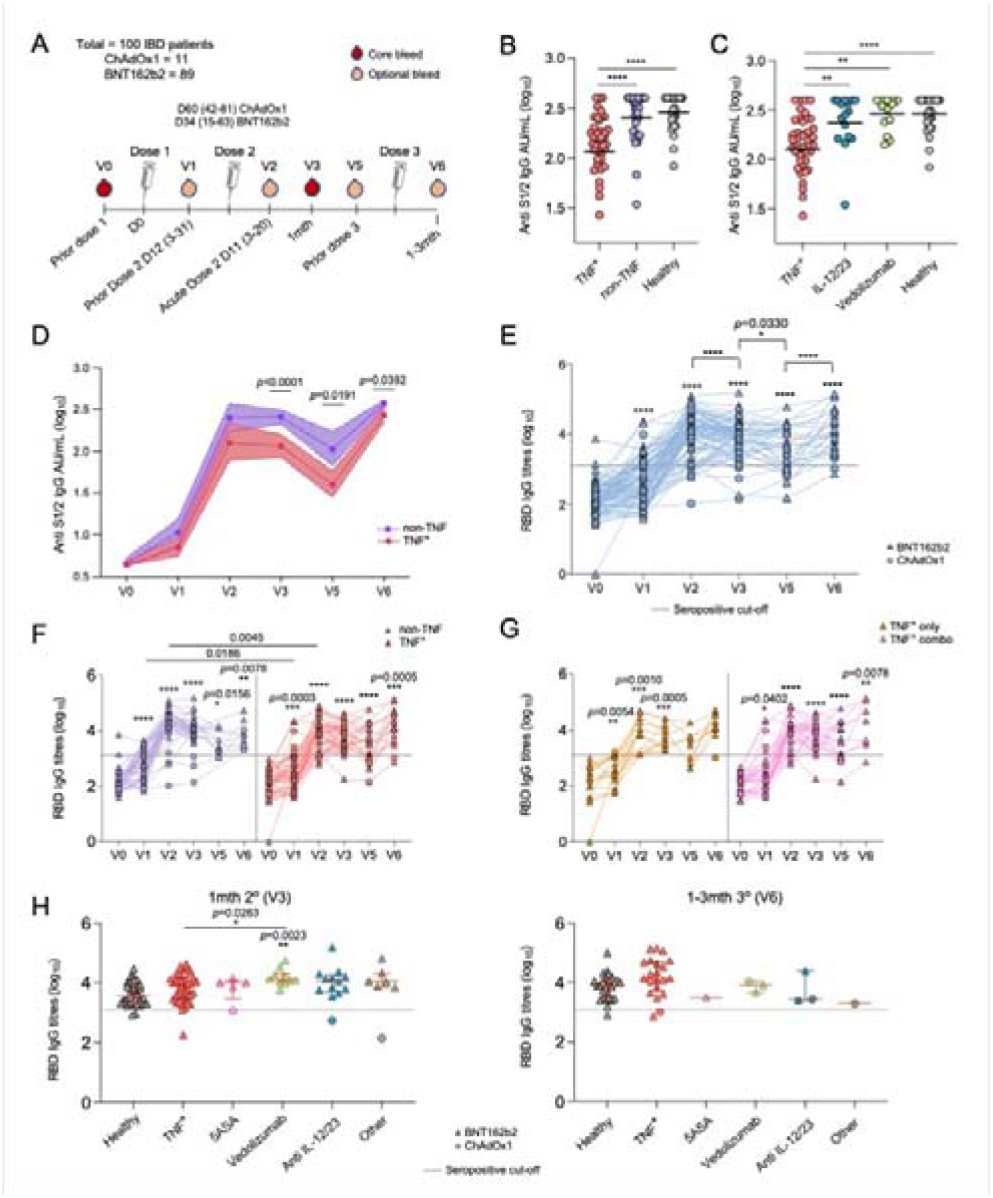
SARS-CoV-2 S1/2- and RBD-specific antibodies in IBD patients after 2 and 3 doses of COVID-19 vaccine. (A) Study cohort and sampling timeline. (B-D) Anti-S1/2-specific antibody titres in (B) IBD patients on anti-TNF or non-TNF treatment versus healthy individuals at ∼1 month post second dose, (C) in TNF, IL-12/IL-23, Vedoluzimab-treated IBD patients versus healthy individuals, and (D) in anti-TNF and non-TNF IBD patients at all time-points. Geometric mean and SD are shown. (E) RBD IgG antibody titres of IBD patients over the course of the study. Seropositive cut-off line is mean+2SD of baseline titres (V0). (F) RBD IgG antibody titres of IBD patients on anti-TNF treatment or non-TNF treatment over the course of the study. (G) RBD IgG titres of IBD patients receiving anti-TNF treatment only or anti-TNF treatment in combination with other therapies. (H) RBD IgG titres of IBD patients across a range of treatments in comparison to healthy controls at V3 and at V6. Statistical significance (two-tailed) was determined by Kruskal-Wallis test between 3 or more groups (B and C), Wilcoxon test for timepoint comparisons against V0 (floating values for E-G), Wilcoxin test between two time-points (connecting lines in E), Mann-Whitney for comparisons between treatment groups at one time-point (D and connecting lines in F) and Dunn’s multiple comparisons set on healthy versus all other disease groups (H). Exact *p* values 0.0001<0.05 are shown except *p*<0.0001=****.

Geometric mean [SD] anti-S1/2 IgG antibody concentrations at ∼1 month after second dose vaccination (V3) were lower in anti-TNF treated patients 126.44 [2.3] compared to non-TNF treated IBD patients 262.1 [1.7] (*p*<0.0001) and healthy controls 295.5 [1.5] (*p*<0.0001) (figure 1B). There was no difference between vedolizumab or IL-12/IL-23-treated patients compared to healthy controls (figure 1c). Out of the 40 patients sampled before and after the third vaccine dose (V5 and V6 timepoints), all participants experienced a rise in anti-S1/2 IgG antibody concentrations following the third vaccine dose (figure 1d). Prior to the third vaccine dose (V5), the mean S1/2 antibody concentration in anti-TNF-treated participants was significantly lower 51.0 [2.6] compared to non-TNF treated participants 116.5 [2.3] (*p*=0.0191, figure 1d). At 4-12 weeks post third dose vaccination (V6), the mean S1/2 antibody concentration in anti-TNF-treated participants, remained significantly lower 285.7 [1.6] compared to non-TNF treated participants 365.3 [1.2] (*p*=0.0392, figure 1d). Most importantly however, these antibody responses after the third vaccine dose were higher than after the second dose.

Sex, type of IBD, smoking, and immunomodulator/methotrexate use did not affect anti-S1/2 IgG antibody concentrations on multivariate analysis (Figure 4). However age over 60 years was associated with significantly lower anti S1/2 titres after second vaccine dose with geometric mean ratio 0.78 (95% CI 0.55-0.96, p = 0.03). Infliximab levels did not influence anti-S1/2 IgG antibody concentration on Pearsons correlation (data not shown).

IgG antibodies against receptor-binding domain (RBD) of ancestral spike protein showed an increase in anti-RBD titres in all 100 IBD patients over the course of the study. RBD IgG titres were significantly higher than baseline (V0) at all timepoints (*p*<0.0001, figure 1e). However, compared to acute responses after the second dose (V2), titres dropped significantly at ∼1 month (V3) and ∼3 months post second dose (V5) (*p*<0.0001 and *p*=0.0330 respectively, figure 1e). Importantly, like the spike-specific IgG antibody responses, RBD IgG titres significantly increased to peak responses after the third dose (V6 versus V5, *p*<0.0001). 4 IBD patients were below the seropositive cut-off at V3, while 2 patients remained seronegative at V6.

IBD patients receiving both anti-TNF and non-TNF treatments showed significant increases in RBD IgG titres at all timepoints post vaccination (figure 1f). Those patients on non-TNF therapies had higher RBD IgG titres at V1 (prior dose 2), V2 (acute post dose 2) and V3 (∼1 month post dose 2) compared to anti-TNF-treated patients, although these differences were diminished after a third dose (figure 1f). Patients who were treated with anti-TNF therapy alone showed comparable antibody responses to those on anti-TNF therapy in combination with immunomodulators (figure 1g). In fact, IBD patients across a range of therapies had very comparable antibody responses when compared to healthy individuals at ∼1 month post second dose (V3) and ∼1-3 months post third dose (V6), with those patients treated with vedolizumab showing higher RBD IgG titres compared to healthy individuals (4.18 vs 3.60 *p*=0.0034) and anti-TNF-treated patients (3.77) at V3 (*p*=0.0263) (figure 1h).

Taken together, three doses of COVID-19 vaccination induced robust SARS-CoV-2-specific antibody responses in IBD patients on anti-TNF and non-TNF based treatments, comparable to healthy individuals.

### COVID-19 vaccination induces strong Tfh activation in the blood of IBD patients

Whole blood analyses of antibody-secreting cells (ASCs), circulating T follicular helper (Tfh) cells, CD4^+^ T cells and CD8^+^ T cells, was performed across V0-V3 to determine activation profiles following the first and second vaccine dose in a subset of 82 IBD patients (figure 2A). ASCs and Tfh cells are transiently activated in the blood ∼7-10 days following COVID-19 mRNA vaccination ^20,21^ as well as following SARS-CoV-2 infection ^18,22^. These cells are typically upregulated in the blood around the same time following influenza vaccination ^23^ and following influenza virus infection ^24^. Here, acute time-points were sampled on average 12 days post first vaccination V1; (range 3-31 days) and 11 days post second vaccination (V2; range 3-20 days).

**Figure 2.**
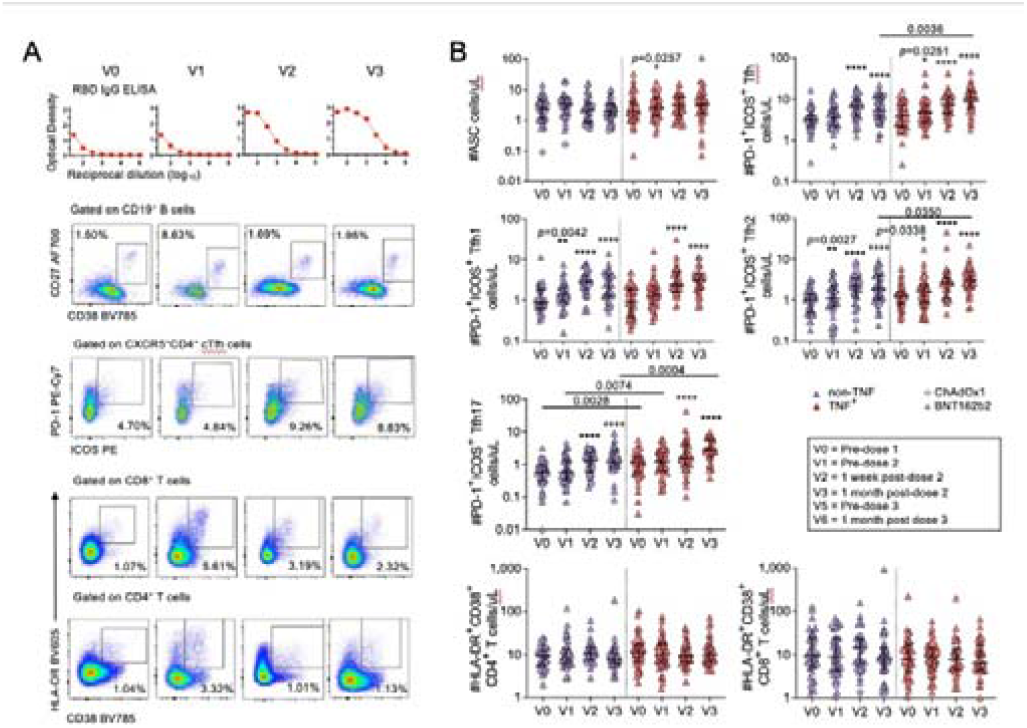
Whole blood analysis of immune cell subsets in the blood of IBD patients before and after COVID-19 vaccination. (A) Representative analysis of 1 IBD patient over V0-V3 timepoints showing RBD titration curves and FACS plots of ASCs and activated Tfh, CD4^+^ and CD8^+^ T cell subsets. (B) Numbers of ASCs, activated Tfh/Tfh1/Tfh2/Tfh17 T cells, and activated CD4^+^ and CD8^+^ T cells in IBD patients on anti-TNF-based or non-TNF based treatment over V0-V3. Statistical significance (two-tailed) was determined by Wilcoxon test for timepoint comparisons against V0 (floating values) and Mann-Whitney for comparisons between treatment groups at one time-point (connecting lines). Exact *p* values 0.0001<0.05 are shown except *p*<0.0001=****.

ASCs increased following the first vaccine dose in anti-TNF-treated IBD patients, but this was not observed acutely after the second dose or at any time-point in non-TNF patients (figure 2b). Circulating CXCR5^+^ Tfh cells from anti-TNF and non-TNF groups were highly activated at the acute timepoint (V2) and ∼1 month following the second dose (V3) by dual expression of PD-1 and ICOS. Increased levels of activation were also observed after the first dose in anti-TNF patients. These observations were evident across the Tfh subsets, including CXCR3^+^CCR6^-^ Tfh1 cells, CXCR3^-^CCR6^-^ Tfh2 cells, and CXCR3^-^CCR6^+^ Tfh17 cells (figure 2b). However, despite the capacity for both treatment groups to induce activated Tfh cell subsets, those patients on anti-TNF therapy displayed significantly higher levels of activated Tfh, Tfh2 and Tfh17 cells at ∼1 month post second dose (V3) compared to non-TNF-treated patients. Non-TNF-treated patients had slightly higher baseline and V1 levels of activated Tfh17 cells (figure 2b). Interestingly, we did not observe significant increases in activation of CD4^+^ and CD8^+^ T cells by dual expression of HLA-DR and CD38, perhaps due to the wide spread of baseline activation levels observed across the IBD patients.

Taken together, we observed highly activated Tfh responses and their subsets in both anti-TNF and non-TNF IBD patients following two doses of COVID-19 vaccination, with some increases after the first dose. Tfh responses are generally less transient than ASCs which decrease rapidly after peaking in numbers. Therefore, although we did not observe an overall prototypical response in ASCs and HLA-DR^+^CD38^+^ T cell subsets, some of the acute and transient peak responses may have been missed by the later sampling time points.

### Robust SARS-CoV-2-specific T cell responses are detected in IBD patients following COVID-19 vaccination

To evaluate SARS-CoV-2-specific T cell responses in IBD patients, we performed an activation induced marker (AIM) assay at baseline (V0), ∼1 month post second dose (V3) and ∼1-3 months post third dose (V6) (figure 3a). PBMC’s were cultured with overlapping spike peptides and evaluated for the upregulation of T cell activation markers. IBD patients showed on average a 10-fold increase in CD134^+^CD137^+^ CD4^+^ T cells and 14-fold increase in CD69^+^CD137^+^ CD8^+^ T cells after the second dose (V3) compared to baseline (V0) (both *p*<0.0001, figure 3b). These responses were maintained after the third dose (V6), which were on average 8-fold and 11-fold higher for CD4^+^ and CD8^+^ T cells, respectively, compared to baseline (*p*=0.0005 and *p*<0.0034, respectively, figure 3b).

**Figure 3.**
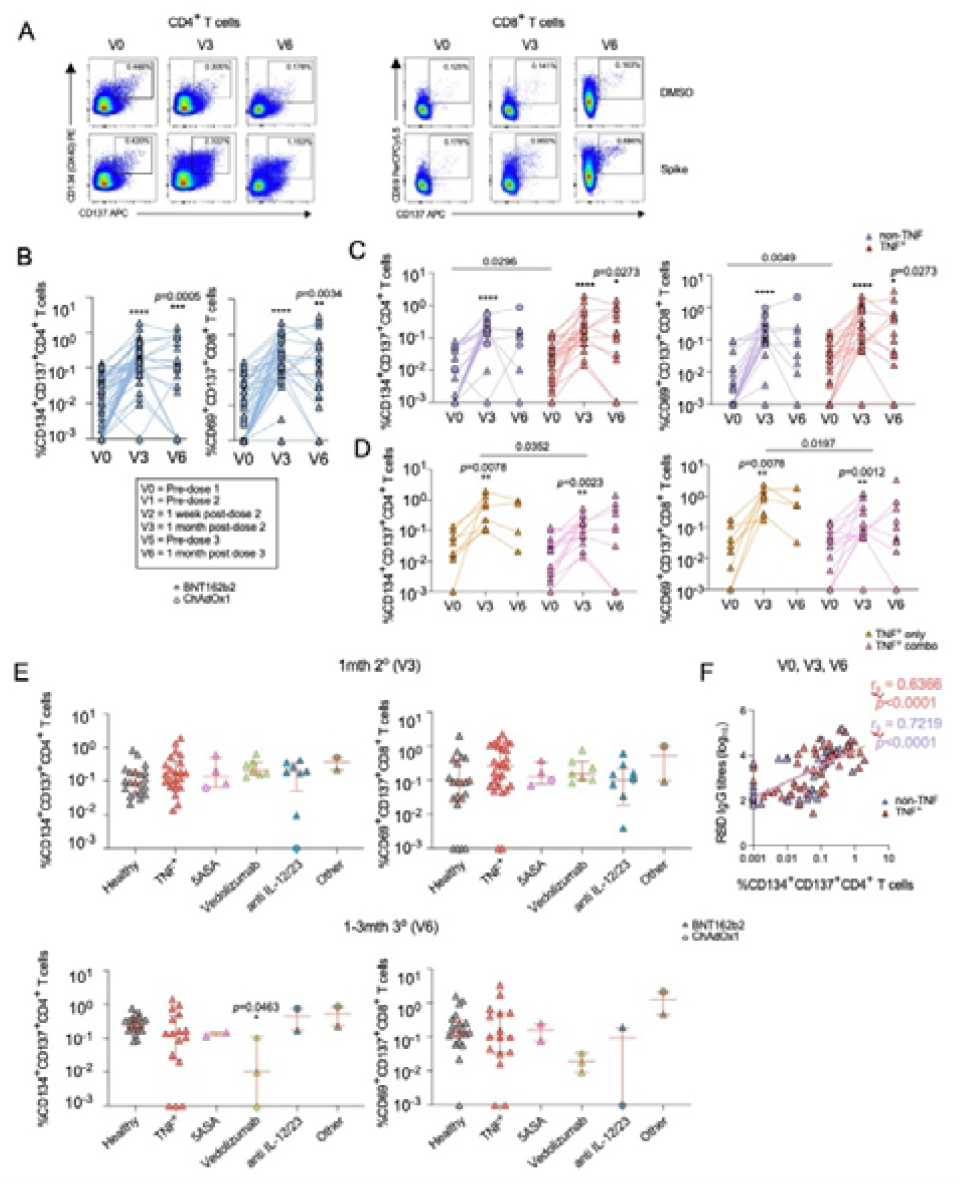
Spike-specific CD4^+^ and CD8^+^ T cell responses in IBD patients before and after COVID-19 vaccination. (A) Representative AIM FACS plots. (B) Frequency of AIM CD4^+^ and CD8^+^ T cells in IBD patients at V0, V3 and V6. (C) AIM CD4^+^ and CD8^+^ T cell responses in IBD patients on anti-TNF-based and non-TNF-based treatments at V0, V3 and V6, and (D) in IBD patients receiving TNF treatment only or anti-TNF treatment in combination with other therapies. (E) AIM CD4^+^ and CD8^+^ T cell responses in IBD patients across a range of treatments in comparison to healthy controls at V3 and V6. (F) Spearman correlation of RBD IgG antibody titres and AIM CD4^+^ T cell responses in IBD patients on anti-TNF-based and non-TNF-based treatments, combining all time-points. Statistical significance (two-tailed) was determined by Wilcoxon test for time-point comparisons against V0 (floating values, B-D), Mann-Whitney for comparisons between treatment groups at one time-point (connecting lines, C-D) and Dunn’s multiple comparisons set on healthy versus all other disease groups (E). Exact *p* values 0.0001<0.05 are shown except *p*<0.0001=****.

**Figure 4:**
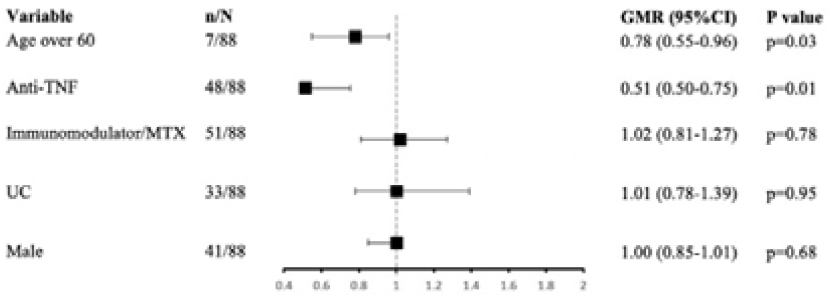
Exponentiated coefficients of a linear regression model of log □□-transformed anti S1/2 antibodies at ∼1 months after second vaccine dose (V3). Values shown are geometric mean ratios of antibody concentrations with each variable.

IBD patients receiving either anti-TNF or non-TNF treatment generated comparable SARS-CoV-2-specific CD4^+^ and CD8^+^ T cell responses following the second vaccine dose (all *p*<0.0001), which were maintained after the third dose (figure 3c). Anti-TNF treated patients did have higher baseline activation of CD4^+^ and CD8^+^ T cells compared to non-TNF patients (V0), but this was not observed following COVID-19 vaccination (*p*=0.0296 and *p*=0.0049, respectively). IBD patients receiving anti-TNF therapy alone or combination anti-TNF therapy both showed significant increases in SARS-CoV-2-specific CD4^+^ (*p*=0.0078 and *p*=0.0023, respectively) and CD8^+^ (*p*=0.0078 and *p*=0.0012, respectively) T cell responses after the second dose (V3), which were maintained after the third dose (V6) (figure 3d). T cell responses after the second dose (V3) were higher for patients receiving anti-TNF alone compared to those receiving combination anti-TNF therapy (*p*=0.0352 for CD4^+^ T cells and p=0.0197 for CD8^+^ T cells), however responses were comparable after the third vaccine dose.

IBD patients across a range of therapies had comparable SARS-CoV-2-specific CD4^+^ and CD8^+^ T cell responses when compared to healthy individuals at ∼1 month post second dose (V3) and ∼1-3 months post third dose (V6), except for patients on vedolizumab, which had lower T cell responses after the third dose, albeit in a small number of patients (n=3). (figure 3e). This contrasts with the vedolizumab group having higher RBD IgG antibody levels after the second dose compared to healthy controls. Nevertheless, RBD IgG antibody titres positively correlated with activated CD134^+^CD137^+^ CD4^+^ T cell responses in both anti-TNF (r_s_=0.6366, *p*<0.0001) and non-TNF patients (r_s_=0.7219, *p*<0.0001), when all time-points were combined (figure 3f).

Overall, comparable to healthy individuals, IBD patients receiving anti-TNF or non-TNF treatment generate a robust SARS-CoV-2-specific CD4^+^ and CD8^+^ T cell response after the second COVID-19 vaccination dose, which is maintained after the third dose.

## DISCUSSION

Patients with IBD are often required to take immune-modifying agents to alleviate disease symptoms with the long-term goal of achieving mucosal healing. We investigated whether these treatments may interfere with the ability of IBD patients to generate prototypical immune responses to SARS-CoV-2 vaccination. Our study provides evidence that IBD patients on either anti-TNF or non-TNF treatment can generate robust and prototypical spike- and RBD-specific antibody responses as well as SARS-CoV-2-specific CD4^+^ and CD8^+^ T cell responses following the second dose, which are maintained ∼1-3 months after the third dose and comparable to healthy individuals. Acute activation of circulating Tfh responses can also be observed after the first and second vaccine dose.

Although IBD patients on anti-TNF therapy had lower spike- and RBD-specific IgG antibody responses after the second dose compared to non-TNF therapy, our study demonstrates that responses were comparable following the third vaccine dose. This contrasts with other studies that have reported antibody concentrations remain attenuated with TNF therapy after third dose vaccination compared to non-TNF treated patients ^8,11,12^. Interestingly, patients receiving vedolizumab had higher RBD IgG antibody levels after the second vaccine dose compared to healthy controls and anti-TNF treated patients, but levels were again at similar levels after the third dose. In our study, 3 of 14 (21%) participants were on dose escalated vedolizumab. Vedolizumab blocks α4β7 expressing lymphocytes, which are unique to the gastrointestinal tract. T regulatory cells suppress virus specific antibody responses ^25^, and hence the impairment of T regulatory cells in the setting of high vedolizumab concentrations ^26^may be a potential mechanism for our finding. Higher antibody responses in vedolizumab patients compared to anti-TNF-treated patients have been reported following the second and third dose ^12^. Despite this, overall, COVID-19 vaccination is highly successful in our cohort of IBD patients receiving various anti-TNF and non-TNF treatments. Out of 100 IBD patients, only 4 patients failed to generate anti-RBD IgG antibody responses after the second dose and only 2 out of 40 patients after the third dose. These results are reassuring in a group considered immunosuppressed. Further, the Surveillance Epidemiology of Coronavirus Under Research Exclusion (SECURE-IBD) registry demonstrates that despite immunosuppression, IBD patients are not at an increased risk of viral acquisition ^27^. Outcomes of COVID-19 infection in IBD patients are also comparable to the general population, although systemic corticosteroids and active IBD are associated with a higher risk of complications ^28,29^.

T cell responses are an integral component of COVID-19 vaccine-induced immunity, which supplements our understanding of the protection and longevity provided by SARS-CoV-2 vaccination. Although T cells do not prevent infection, they excel in the clearance of intracellular pathogens through recognition and lysis of virally-infected cells, activating macrophages and support B cell maturation by way of Tfh cells ^16^. Most published data focus on vaccine-induced humoral responses, which has become less efficacious against the dominant circulating Omicron virus variants. This is because the original vaccines were targeted at the spike protein of the ancestral SARS-CoV-2 variant, while Omicron variants possess over 30 mutations in the spike region ^30,31^. Hence neutralising antibody activity in recipients of 2 doses of the BNT162b2 vaccine is estimated to be reduced by 10-fold, with an even greater reduction in recipients of 2 doses of the ChAdOx1 vaccine, compared to the Delta variant. Despite the diminished neutralising activity of vaccine-induced humoral responses, T cells retain efficacy against mutated viral variants as they are directed against multiple targets including membrane (M) and nucleocapsid (N), which are preserved ^17^. In our study, we observed an increase in activation of Tfh cells following first and second vaccine dose in both anti-TNF and non-TNF IBD patient groups. We also observed durable and comparable SARS-CoV-2-specific CD4^+^ and CD8^+^ T cell responses after second and third doses, where CD4^+^ T cell responses correlated with RBD IgG antibody responses in both treatment groups.

Although infection with Omicron variants result in milder symptoms, vaccination remains an important consideration as higher transmissibility and immune escape prompting re-infection within a shorter interval in recovered patients places a significant burden on health systems worldwide. In the CLARITY study, infliximab treated patients were two-fold more likely to contract SARS-CoV-2 infection compared to vedolizumab treated patients ^12^ in the Omicron era. T cell responses are even more crucial in groups with attenuated antibody responses. Reassuringly in anti-TNF-treated patients who experience a more rapid decline in humoral protection with time, T cell responses are not impaired and in fact augmented with vaccination ^32^. The biological mechanism behind this is not clear. This, in addition to role of anti-TNFs in abrogating the cytokine storm associated with severe COVID-19 infection, may underpin the finding that anti-TNF monotherapy is shown to be associated with reduced odds of hospitalisation and death ^29,33^. Hence a durable T cell response may provide some protection against severe SARS-CoV-2 infection in those with waning antibody titers or who do not mount a strong antibody response.

Although vaccine efficacy is impaired against symptomatic SARS-CoV-2 infection of viral variants, vaccination continues to provide protection against severe disease, hospitalisation and death ^34^. Furthermore, vaccines may minimise the interruption to IBD treatment that severe, or prolonged SARS-CoV-2 infection may pose in IBD patients. Misconceptions^35^ and complacency may hinder vaccine uptake, however boosters should continue to be encouraged. This study supports vaccination in IBD patients on multiple therapies and demonstrates reassuring immune responses to vaccination. The introduction of bivalent booster vaccines will also enhance this effect ^36^.

The main limitation of this study was the small samples size of IBD patients within each treatment group, however we recruited a larger cohort of 100 patients to represent a heterogenous population of patients on different treatments. We were unable to analyse the influence of tofacitinib due to small numbers. The strengths of our study were the ability to assess the breadth of both humoral and cellular responses at baseline before vaccination, prior to being exposed to SARS-CoV-2, and across clearly defined timepoints after vaccination with a healthy control arm.

In conclusion, anti-TNF therapy slightly impairs the humoral response to SARS-COV-2 vaccination which can be boosted with additional vaccine doses. However, overall, IBD patients displayed similar frequencies of SARS-CoV-2-specific CD4^+^ and CD8^+^ T cells compared to healthy vaccinated controls which was not impacted by anti-TNF therapy. In the setting of immune escape with viral variants, the cellular response will play an increasingly important role in the protection against severe SARS-CoV-2 infection.

## Supporting information

STROBE

## Data Availability

All data produced in the present study are available upon reasonable request to the authors

## Acknowledgments

We thank Beverly Cox, Meryem Fezollari and Elizabeth Shilling (Royal Melbourne Hospital) for support with the healthy vaccinated cohort; Professor Florian Krammer (Icahn School of Medicine at Mount Sinai, New York) for ancestral RBD protein; and BEI Resources, NIAID, NIH for providing Peptide Array, SARS-Related Coronavirus 2 Spike (S) Glycoprotein, NR-52402.

